# Changes in public perception of AI in healthcare after exposure to ChatGPT

**DOI:** 10.1101/2025.01.23.25321048

**Authors:** Anders Aasted Isaksen, Jonas Rosborg Schaarup, Lasse Bjerg, Adam Hulman

**Affiliations:** Steno Diabetes Center Aarhus, Aarhus University Hospital, Denmark; Department of Public Health, Aarhus University, Denmark

## Abstract

**Background:** Artificial intelligence (AI) is expected to become an integral part of healthcare services, and the widespread adoption of AI tools in all areas of life is making AI accessible to the general public. Public perception of the benefits and risks of AI in healthcare is key to large-scale acceptance and implementation, and is increasingly influenced by first-hand experiences of AI. The aim of this study was to assess how exposure to ChatGPT changed public perception of AI in healthcare.

**Methods:** We used baseline and follow-up data from 5,899 survey participants, who reported their perception of AI in 2022 and 2024, and ChatGPT use in 2024. Administrative and healthcare data from nationwide Danish registers was used for weighting and adjustment. Multinomial multivariate logistic regression was used to model how exposure to ChatGPT use affected changes in perception of AI.

**Results:** At baseline (before ChatGPT’s launch) 2,236 individuals (37%) were unsure of the benefits and risks of AI in healthcare, 2,384 (40%) perceived net benefits, 1,083 (18%) perceived benefits and risks as equal, and 196 (3.3%) perceived net risks. At follow-up, 1,195 individuals (20%) had been exposed to ChatGPT use, which was associated with higher odds of changing perception of AI to benefits (OR 3.21 [95% CI: 2.34-4.40]) among individuals who were unsure at baseline, and lower odds of changing to uncertainty from more defined baseline perceptions (from benefits (OR 0.32 [0.24-0.42]), equal (OR 0.47 [0.32-0.69]) and risks (OR 0.27 [0.08-0.98])).

**Conclusion:** Exposure to ChatGPT was associated with a change towards positive perception of benefits and risks of AI in healthcare among individuals who were uncertain prior to exposure, and individuals with more defined perceptions of AI were less likely to become uncertain after exposure to ChatGPT.

## Background

The implementation of artificial intelligence (AI) technology in healthcare is expected to increase efficiency, improve care quality, relieve the growing burden on healthcare systems,^1^ and empower patients’ access to knowledge.^2^ The practical implementation of this technology is growing at a rapid pace, as evidenced by the number of annual Food and Drug Administration (FDA) approvals for AI-enabled medical devices, which increased from around five to a thousand in the last decade.^3,4^ With the prospect of AI being widely implemented in healthcare settings, exposing patients and the general public to these tools, it is important to understand how first-hand experiences with AI tools influence perceptions of benefits and risks of applying AI in healthcare - particularly since a large part of the public is uncertain or undecided about their perception of AI.^5–7^

Previously, one of the challenges for the public to form a clear opinion on the application of AI was that these technologies were invisible elements of daily life.^8^ In recent years, AI has become a visible, accessible and practical tool that the public increasingly uses and interacts with in everyday life - particularly since the launch of ChatGPT in November 2022 and its rapid adoption, sometimes referred to as the “large language model revolution”.^9^ With this change, the public’s perception of AI is likely to be shaped by practical experiences from first-hand use rather than intangible or theoretical considerations. Patients were quick to adopt ChatGPT and other general-purpose generative AI tools for guidance on personal healthcare conditions, highlighting the close relationship between AI and healthcare.^10^ While the public’s overall perception of AI in healthcare has been studied in cross-sectional surveys across the world,^5,6,11^ the evidence on the impact of first-hand exposure to AI and longitudinal changes in perception is sparse.

During discussions with members of the user panel of patients and relatives at Steno Diabetes Center Aarhus, a diabetes outpatient centre in Denmark,^12^ about the results of a previous survey of perception of AI in healthcare,^5^ some members noted how impressed they were with ChatGPT and remarked that they had changed their perception after their first use. Inspired by this, we hypothesized that ChatGPT exposure would most strongly impact the large group of the public (one-third of survey respondents) who were unsure about AI’s benefits and risks. This study aims to provide evidence on the impact of being exposed to first-hand use of AI tools, such as ChatGPT, on the perception of AI in healthcare. Specifically, we investigate changes in public perception of AI in healthcare between 2022 and 2024, and assess the impact of exposure to ChatGPT on change in perception from the baseline perception reported before ChatGPT’s release.

## Materials and Methods

### Data collection

In this study, we collected data on perception of AI and ChatGPT use, and combined it with administrative data from nationwide Danish registers.

Survey data: Health in Central Denmark (HICD) is a digital and postal questionnaire survey initially conducted in 2020 on all inhabitants of Central Denmark Region aged 18 to 74 years identified as prevalent diabetes cases in register data on 31 December 2018, plus an equally-sized group of people without diabetes (matched to diabetes cases by sex, age, and municipality)^13^. Among respondents in 2020 that volunteered to be contacted for further studies, a baseline survey on perception of AI was conducted in the spring of 2022,^14^ and these individuals were contacted again in 2024 as part of the next wave of the HICD survey, which provided follow-up data for this study.^15^

Perceived risks and benefits of AI in healthcare were assessed in baseline (2022) and follow-up (2024) surveys. Participants were asked, “Do you think the benefits of using artificial intelligence in the healthcare sector outweigh the risks?” with four answers: “Benefits outweigh risks” (“Benefits”), “Benefits equal risks” (“Equal”), “Risks outweigh benefits” (“Risks”), and “Don’t know” (the 2024 questionnaire contained two “don’t know”-responses options that were merged for this study).^15^ In the 2022 questionnaire, participants who reported having never heard of AI were not subsequently asked about their perception, but were classified as “Don’t know”. Information on exposure to ChatGPT use was reported in 2024 with the question “Have you used ChatGPT?” with the following answer options: “Yes, I use it often”, “Yes, I have tried it”, “No, but I have heard about it” or “No, I have not heard about it”. In the analyses, this variable was included as binary: “ChatGPT exposed” and “Unexposed”, by merging the two former and latter categories, respectively.

Administrative and healthcare data covering the entire Danish population can be linked at the individual level.^16^ Information on age, sex, and education level (defined by the total length of attained of education) was obtained from the Danish Civil Registration System^17^ and the Danish Population Education Register.^18^ Diabetes status was defined as prevalent “Type 1 diabetes” (T1D), “Type 2 diabetes” (T2D), or “No diabetes” at baseline according to the Open-Source Diabetes Classifier based on HbA1c measurements ≥48 mmol/mol (6.5%), hospital diagnoses of diabetes, diabetes-specific podiatrist services, and purchases of glucose-lowering drugs.^19^

### Statistical Analysis

The distribution of study population characteristics was tabulated by exposure to ChatGPT use and reported as frequencies (%) or mean (SD). Alluvial plots and cross-tabulations were applied to visualize raw changes in perceptions of AI from baseline to follow-up between ChatGPT exposed and unexposed individuals. Multinomial logistic regression was used to model the association between ChatGPT use and the perception of AI use in healthcare in the follow-up survey, stratified by perception of AI in the baseline survey. To assess the change in perception from baseline, the reference category of the outcome differed between each strata to reflect the baseline perception in each group. The models were adjusted for age (continuous), sex, education level (“<10 years”, “10-15 years”, and “>15 years”) and diabetes status and applied stabilized propensity score weights based on the distribution of these covariates in the background population (capped at 95% percentile). Supplementary analyses include unweighted analyses and analyses using weights computed against the 90,854 invitees in the initial 2020 HICD survey (Figure S3 and S4), and overall analyses using unstratified models adjusted for baseline perception (Table S6). Statistical analyses were pre-registered^20^ and performed using R version 4.4.1^21^ and packages for data processing,^22^ weighting,^23^ regression modelling,^24^ and visualizations.^25,26^

## Results

The flow of participants from the background population into the study population is shown in Figure 1. Of 90,854 individuals invited to the initial HICD survey, 12,755 (14%) consented to be contacted for further studies, of whom 5,934 (47%) responded in both the 2022 and 2024 surveys. After excluding 35 individuals without information on education level, the study population consisted of 5,899 individuals, of whom 365 (6.2%) had T1D and 2,540 (43%) had T2D.

**Figure 1:**
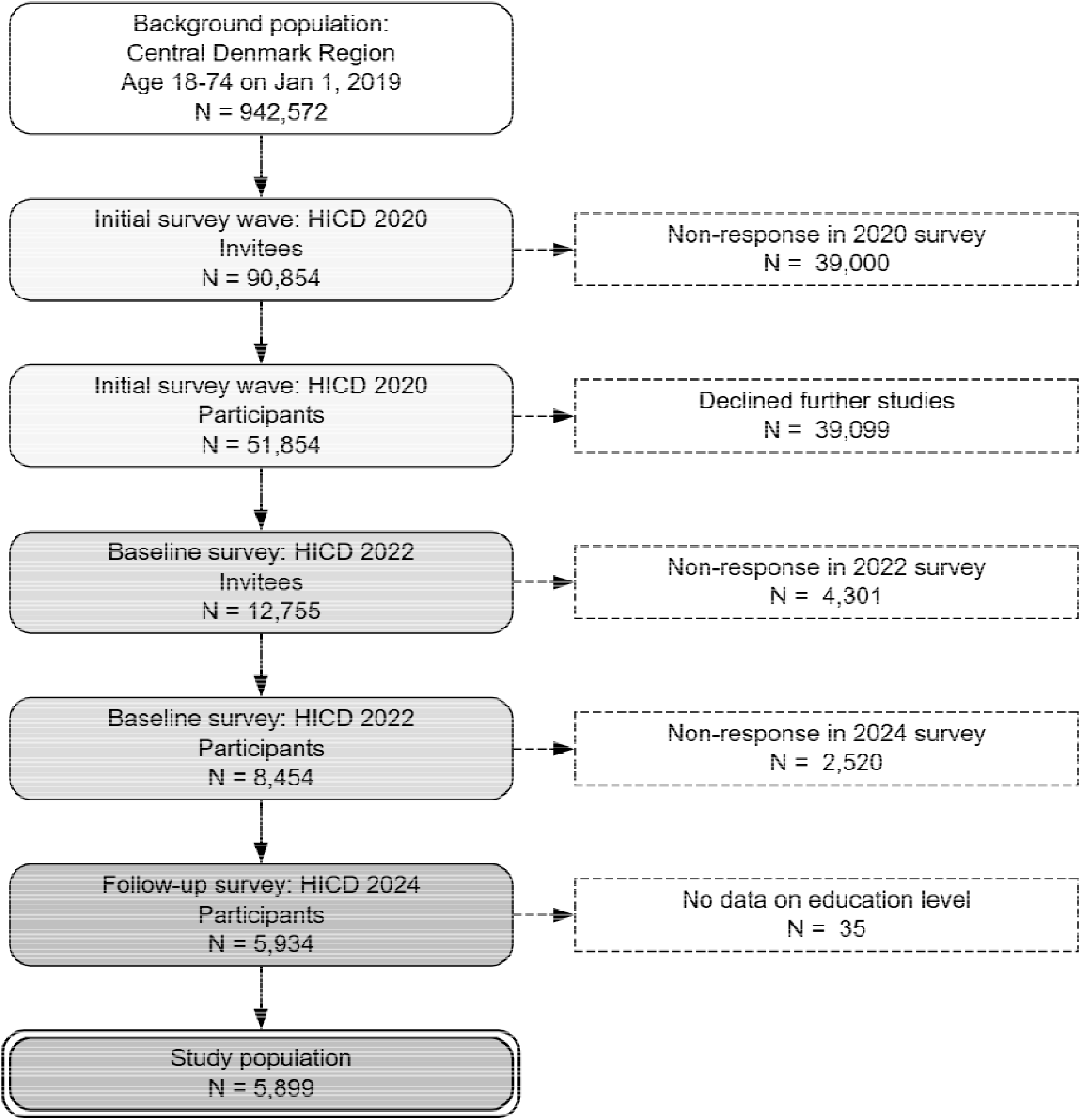
Flow of individuals from the background population of the Central Denmark Region into the two waves of the Health In Central Denmark (HICD) survey and the study population.

Overall characteristics of the study population and by ChatGPT exposure are shown in Table 1. At baseline, 2,236 of these 5,899 individuals (37%) were unsure of the benefits and risks of using AI in healthcare, while 2,384 (40%) perceived the benefits as greater than the risks, 1,083 (18%) perceived the benefits as equal to the risks, and 196 (3.3%) perceived the risks as outweighing the benefits.

**Table 1:** Distribution of baseline characteristics and perceptions of benefits and risks of AI in healthcare at baseline and at follow-up in the study population and among individuals exposed to ChatGPT use or unexposed.

|  |  | Overall | ChatGPT exposed | No ChatGPT exposure |
| --- | --- | --- | --- | --- |
| Total N (%) |  | 5,899 | 1,195 (20.3) | 4,704 (79.7) |
| Sex | Men | 3,649 (61.9) | 824 (69.0) | 2,825 (60.1) |
| Age (years) | Mean (SD) | 65.4 (8.9) | 61.4 (10.2) | 66.4 (8.2) |
| Diabetes status | No diabetes | 2,994 (50.8) | 647 (54.1) | 2,347 (49.9) |
|  | Type 1 diabetes | 365 (6.2) | 87 (7.3) | 278 (5.9) |
|  | Type 2 diabetes | 2,540 (43.1) | 461 (38.6) | 2,079 (44.2) |
| Education level (years) | < 10 | 289 (4.9) | 17 (1.4) | 272 (5.8) |
|  | 10 - 15 | 3,334 (56.5) | 530 (44.4) | 2,804 (59.6) |
|  | > 15 | 2,276 (38.6) | 648 (54.2) | 1,628 (34.6) |
| Perception of AI at baseline | Don't know | 2,236 (37.9) | 226 (18.9) | 2,010 (42.7) |
|  | Benefits | 2,384 (40.4) | 735 (61.5) | 1,649 (35.1) |
|  | Equal | 1,083 (18.4) | 202 (16.9) | 881 (18.7) |
|  | Risks | 196 (3.3) | 32 (2.7) | 164 (3.5) |
| Perception of AI at follow-up | Don't know | 2325 (39.4) | 205 (17.2) | 2120 (45.1) |
|  | Benefits | 2044 (34.6) | 666 (55.7) | 1378 (29.3) |
|  | Equal | 1132 (19.2) | 238 (19.9) | 894 (19.0) |
|  | Risks | 398 (6.7) | 86 (7.2) | 312 (6.6) |

At follow-up, 1,195 individuals (20%) had been exposed to ChatGPT use. These individuals tended to be younger (mean age 61 years vs. 66 years among unexposed), had a higher proportion of men (69% vs. 60%), lower proportion with diabetes (46% vs. 50%), and had a higher education (54% vs. 35%) than those unexposed. Only 155 (13%) of the 1,195 ChatGPT-exposed individuals reported using ChatGPT often, while 1,223 (26%) of the 4,704 unexposed individuals reported having never heard about ChatGPT.

At baseline, individuals who were subsequently exposed to ChatGPT were more likely to perceive benefits of AI in healthcare (62% vs. 35%) and less likely to be unsure (19% vs. 43%) than unexposed, while the two groups differed less in terms of proportions of individuals reporting “Equal” (17% vs. 19%) or “Risks” (2.7% vs. 3.5%). Study population characteristics weighted against the background population are available in Table S1, and characteristics stratified by baseline perception are available in Table S2 of the Supplementary Appendix.

In the follow-up survey, the overall proportion of individuals reporting “Benefits” decreased to 35%, while the proportion perceiving “Risks” doubled to 6.7% (p < 0.0001). Among individuals who reported “Don’t know” at baseline, 65 (29%) of the 226 who were subsequently exposed to ChatGPT use changed perception to “Benefits”, compared to 272 (14%) of the 2,010 unexposed individuals from this baseline group (Figure 2).

**Figure 2.**
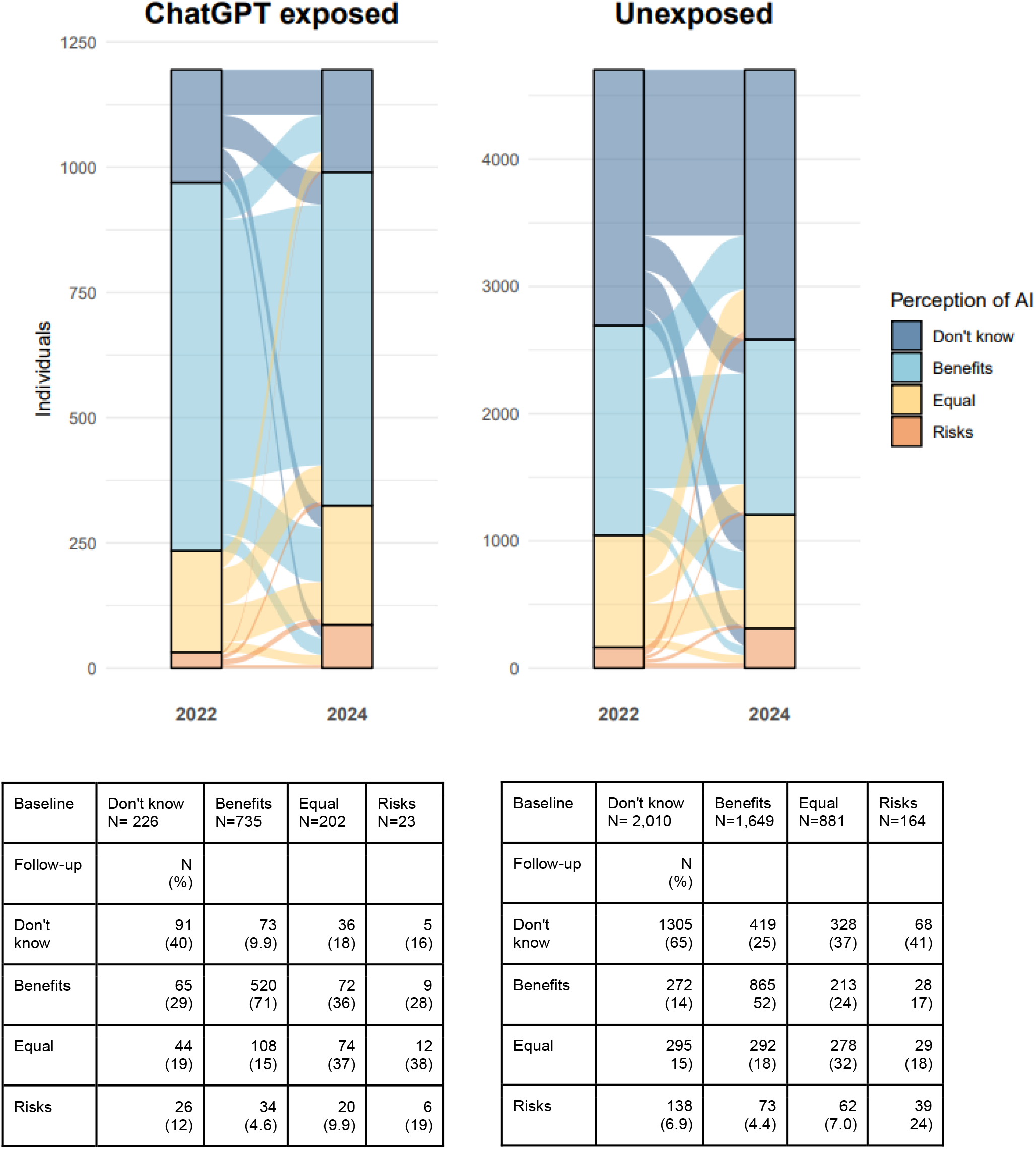
Changes in distribution of perception from baseline (2022) to follow-up (2024) among individuals exposed to ChatGPT use (left) and those unexposed (right). Note the differing Y-axes.

Figure 3 presents the odds ratios (ORs) for changing perception versus maintaining baseline perception at follow-up associated with exposure to ChatGPT use (e.g. an OR > 1 indicates a higher likelihood of change in perception among individuals exposed to ChatGPT compared to unexposed individuals) for each baseline perception group. Exposure to ChatGPT use was associated with higher odds of changing perception to “Benefits” (OR 3.21 [95% CI: 2.34-4.40]) or to “Equal” (OR 1.46 [1.01-2.11]) vs. maintaining the baseline perception in individuals who reported “Don’t know” at baseline. In the remaining baseline perception groups, exposure to ChatGPT use was associated with lower odds of changing perception to “Don’t know” vs. maintaining the baseline perception at follow-up among individuals reporting “Benefits” (OR 0.32 [0.24-0.42]), “Equal” (OR 0.47 [0.32-0.69]) or “Risks” (OR 0.27 [0.08-0.98]) at baseline. A full list of model coefficients is available in supplementary (Table S5).

**Figure 3.**
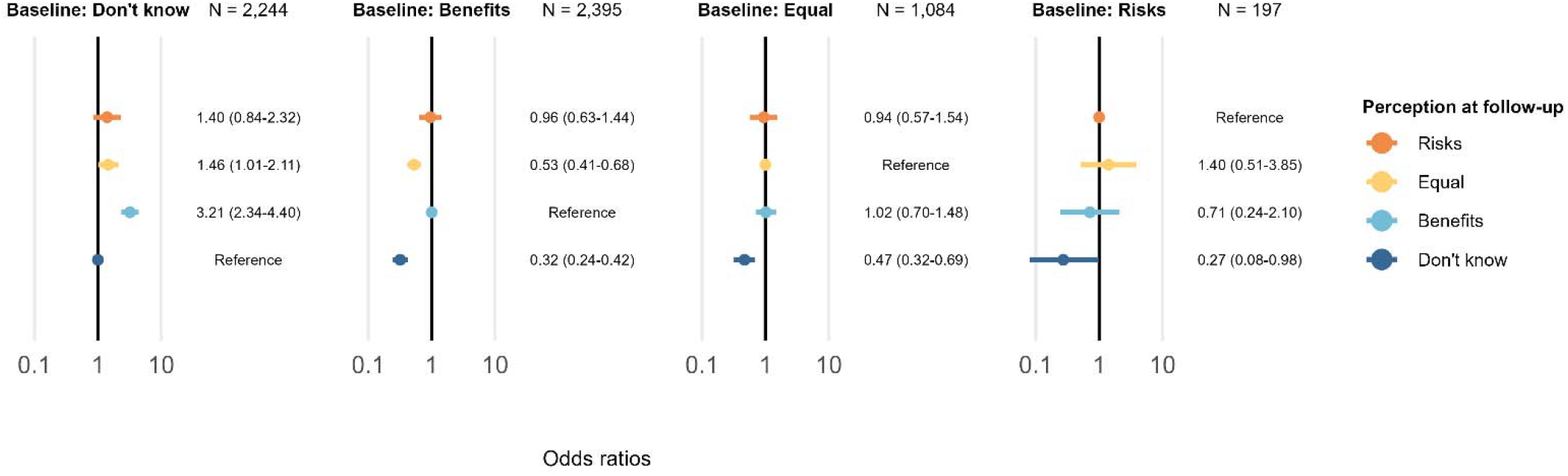
Odds ratios of changes in perception from baseline to follow-up associated with exposure to ChatGPT use stratified by baseline perception from a multinomial logistic regression model adjusted for age, sex, education level, and diabetes status, and weighted with stabilized propensity score weights based on the distributions of these covariates in the background population. Note that the outcome reference category differs between the strata to match the baseline perception level.

In supplementary analyses (Figure S3 & S4) using unweighted models and models weighted to the characteristics of invitees to the initial HICD survey, the estimates for ChatGPT exposure were consistent with the main analysis. However, the estimates for change of perception from “Don’t know” at baseline towards “Risks” (OR 2.35 [1.45-3.81]), “Equal” (OR 1.84 [1.25-2.73]) and “Benefit” (OR 2.92 [2.04-4.17]) at follow-up differed in magnitude between the unweighted and the main analyses.

Some baseline characteristics were associated with change of perception at follow-up in the regression models (Table S5). Generally, shorter length of education, older age and female sex were associated with lower odds of changing baseline perception from “Don’t know” to a more defined perception at follow-up - especially to “Benefits”, and these groups were also more likely to change perception to “Don’t know” at follow-up from other baseline perceptions. In overall analyses adjusted for - rather than stratified by - baseline perception, exposure to ChatGPT use was associated with lower odds of reporting “Don’t know”, “Equal” or “Risks” than “Benefits” at follow-up (Table S6).

## Discussion

In this study, we found that exposure to ChatGPT reduced uncertainty of perceived benefits and risks of AI applied in healthcare, regardless of perception of AI prior to ChatGPT exposure. Furthermore, we found that exposure to ChatGPT mainly resulted in a change towards a positive perception of AI in healthcare among individuals who were uncertain prior to this exposure. Overall, the perception of AI in healthcare became more negative between 2022 and 2024, with a substantial decrease in the proportion of individuals perceiving mainly benefits and more than a doubling of those perceiving mainly risks. While these trends were present in both the ChatGPT exposed and unexposed group, it was less pronounced among individuals exposed to ChatGPT.

Overall, exposure to ChatGPT use was associated with shifts toward more positive perceptions of AI compared to non-exposure. This was driven by shifts from uncertainty at baseline to positive perception at follow-up, without any evidence of changes toward more positive or skeptic perceptions in individuals with other baseline perceptions. Unweighted analyses showed a more diverse influence of ChatGPT exposure in the uncertain group, with higher likelihood of shifting towards both risks and benefits. The study population was older than the background population due to the HICD survey invites being sampled based on the age distribution of individuals with diabetes, and this difference between the weighted and unweighted analyses may be due to a different influence of ChatGPT use among older uncertain individuals than among younger.

Uptake of this technology in the public was limited, as only one-in-five survey respondents had tried ChatGPT (and one-in-five had not even heard of it). Uptake was particularly poor among people with shorter education, women, and older age groups - characteristics which were all associated with changes towards negative or uncertain perceptions of AI or maintaining these skeptic perceptions from baseline to follow-up. This finding adds to previous reports of more sceptical outlooks on AI among individuals with low digital familiarity in the United Kingdom (UK).^27^ Thus, improving digital literacy in these demographics may reduce uncertainty and skepticism regarding benefits and risks of AI in healthcare.

As current and future patients, the public is a key stakeholder in the the implementation of AI in the healthcare system, and their perception of the benefits and risks of the technology may be critical to large-scale implementation and adoption of this technology.^28^ Public perception of the technology remained largely positive, with only a small minority perceiving risks as outweighing benefits at follow-up, but the growing skepticism towards AI in healthcare may become a concern if this trend grows in the coming years. Our study suggests that exposure to AI tools like ChatGPT influenced public perception of AI and could help mitigate growing skepticism, particularly as nearly half of the ChatGPT-unexposed group was undecided at follow-up.

Our study demonstrates the potential for changing public perceptions of AI in healthcare by reducing uncertainty among individuals through exposure to AI tools like ChatGPT. However, negative experiences with poorly performing AI tools could shape perceptions in a negative direction, and evidence on this dynamic remains sparse (File S7). To our knowledge, this is the first study to report how exposure to an AI tool directly impacts public perceptions of AI in healthcare, as the few existing studies on changes in perception after exposure to AI were limited to specific groups, such as healthcare professionals or patients. For instance, emergency radiology professionals reported reduced uncertainty and increased perceived benefits of AI following its implementation,^29^ while urolithiasis patients exposed to ChatGPT-generated notes experienced a negative shift in attitudes.^30^ Thus, there is a lack of evidence to guide the implementation of AI in healthcare, which is seeing increasing uptake across areas of healthcare and data modalities, such as electronic health record analysis for clinical diagnostics and prediction,^31^ clinical image classification,^32,33^ and voice recordings of patients for out-of-hours cardiac arrest detection^34^ and ambient AI scribes.^35^

We observed a growing skepticism toward AI in healthcare from 2022 to 2024 in our study, similar to trends in the UK’s Public Attitudes to Data and AI Tracker Survey, which found that the public became more pessimistic about the impact of AI on society from 2021 to 2023, partly due to data and privacy concerns.^27^ These findings contrast a previous study based on data from social media, which found a positive trend in sentiment towards AI in medical imaging over time from 2019 to 2024^36^. This inconsistency may be due to differences in methodology and settings, but may also reflect that the public’s perception of AI is subject to external factors such as media coverage of AI and digital technology. The sentiment of media coverage may change substantially over time, geography and media source. Within research on the medical knowledge of ChatGPT, several reports have praised it for matching or exceeding the quality of human doctors in medical licensing exams^37–40^ and for showing higher levels of empathy in written communication with patients.^41^ In contrast, other studies have highlighted how ChatGPT may perform significantly worse than human doctors in more complicated, free-text clinical case exams,^42^ and how minor nuances in the wording of inputs can lead to highly biased outputs.^43^ Depending on which of these messages gets picked up by the latest mainstream media coverage, the public perception may shift according to this, rather than the public’s first-hand experiences with AI.

The strengths of this study include the large study population, which allowed for stratified analyses to identify specific changes rather than an overall effect associated with exposure to ChatGPT use. As the baseline perception data was collected shortly before the launch of ChatGPT, this allowed us to closely study the changes in perception after exposure to ChatGPT. Finally, the population-wide register data enabled weighted analyses to increase the representativeness of results and reduce the risk of selection bias inherent in survey-based studies.

There are some limitations to the study. Due to the observational nature of the study, confounding cannot be ruled out. As we did not assess all potential confounders of change in perception of AI that may have been related to personal use of ChatGPT, e.g. exposure to other AI tools or awareness of positive or negative media reports on use of AI or digital technology in healthcare, we cannot rule out that such external factors biased our results. Also, while the item on perception of AI in healthcare was similar in the baseline and follow-up surveys, the remaining content of the questionnaires differed, which could bias responses.^44^ Another limitation of this study is that the population represented the general public, who likely use ChatGPT as a general-purpose tool rather than for healthcare-specific purposes. While diabetes status did not influence the results, the findings may not apply to all patient populations or clinical AI use cases. Public perception of healthcare-specific AI tools likely depends on the tool, its user experience, and its context - factors beyond the scope of this study.

## Conclusion

Exposure to ChatGPT was associated with a change towards positive perception of benefits and risks of AI in healthcare among individuals who were uncertain prior to exposure. Individuals with more defined perceptions of AI prior to exposure to ChatGPT were not swayed towards positive nor negative perceptions, but were less likely to become uncertain after exposure to ChatGPT. Public acceptance of AI in healthcare is key to large-scale implementation of the technology, and while we found increasing skepticism towards AI in healthcare among the public over time, this study demonstrates the potential for changing the public perception of AI in healthcare through exposure of the public to AI tools.

## Data Availability

The Health in Central Denmark data is hosted on remote servers at Statistics Denmark. The project is managed by a steering committee at Steno Diabetes Center Aarhus, Aarhus University Hospital. The steering committee encourages interested researchers to use this resource. More information is available on the website: https://www.stenoaarhus.dk/research/resources/health-in-central-denmark/

https://figshare.com/articles/online_resource/Patients_perception_of_digital_technologies_and_artificial_intelligence_an_online_survey/19248214/1?file=34202812

https://figshare.com/articles/online_resource/Peoples_perception_of_artificial_intelligence_and_chatbots_an_online_survey_from_Health_in_Central_Denmark/27894180/2

## Notes

### Competing Interest Statement

AAI, JRS, LB, and AH are employed at Steno Diabetes Center Aarhus which is partly funded by an unrestricted grant from the Novo Nordisk Foundation (NNF17SA0031230). AAI and AH are supported by a Data Science Emerging Investigator grant (NNF22OC0076725) by the Novo Nordisk Foundation. The funders had no role in the design of the study.

### Clinical Protocols

https://figshare.com/articles/online_resource/_/26048662?file=47115853

### Funding Statement

This study did not receive any funding.

### Author Declarations

The IRB of the Health In Central Denmark Study at Steno Diabetes Center Aarhus gave ethical approval for this work.

## References

1. Topol EJ. High-performance medicine: the convergence of human and artificial intelligence. Nat Med. 2019;25(1):44–56. doi:10.1038/s41591-018-0300-7

2. Rotenberg D, Clark M, Bailey S. Chatbots in Health Care: Connecting Patients to Information: Emerging Health Technologies. Canadian Agency for Drugs and Technologies in Health; 2024. Accessed December 16, 2024. http://www.ncbi.nlm.nih.gov/books/NBK602381/

3. Muralidharan V, Adewale BA, Huang CJ, et al. A scoping review of reporting gaps in FDA-approved AI medical devices. NPJ Digit Med. 2024;7:273. doi:10.1038/s41746-024-01270-x

4. Health C for D and R. Artificial Intelligence and Machine Learning (AI/ML)-Enabled Medical Devices. FDA. Published online July 8, 2024. Accessed December 16, 2024. https://www.fda.gov/medical-devices/software-medical-device-samd/artificial-intelligence-and-machine-learning-aiml-enabled-medical-devices

5. Schaarup JFR, Aggarwal R, Dalsgaard EM, et al. Perception of artificial intelligence-based solutions in healthcare among people with and without diabetes: A cross-sectional survey from the health in Central Denmark cohort. Diabetes Epidemiol Manag. 2023;9:100114. doi:10.1016/j.deman.2022.100114

6. Scantamburlo T, Cortés A, Foffano F, et al. Artificial Intelligence across Europe: A Study on Awareness, Attitude and Trust. IEEE Trans Artif Intell. Published online 2024:1–14. doi:10.1109/TAI.2024.3461633

7. Zhang B, Dafoe A. Artificial Intelligence: American Attitudes and Trends. Published online January 9, 2019. doi:10.2139/ssrn.3312874

8. Yigitcanlar T, Degirmenci K, Inkinen T. Drivers behind the public perception of artificial intelligence: insights from major Australian cities. AI Soc. 2024;39(3):833–853. doi:10.1007/s00146-022-01566-0

9. Mesko B. The ChatGPT (Generative Artificial Intelligence) Revolution Has Made Artificial Intelligence Approachable for Medical Professionals. J Med Internet Res. 2023;25(1):e48392. doi:10.2196/48392

10. Goldberg C. Patient Portal — When Patients Take AI into Their Own Hands. NEJM AI. 2024;1(5):AIp2400283. doi:10.1056/AIp2400283

11. Witkowski K, Okhai R, Neely SR. Public perceptions of artificial intelligence in healthcare: ethical concerns and opportunities for patient-centered care. BMC Med Ethics. 2024;25(1):74. doi:10.1186/s12910-024-01066-4

12. About Steno Diabetes Center Aarhus. Steno Diabetes Center Aarhus - for fagfolk. Accessed January 19, 2025. https://www.stenoaarhus.dk/english/

13. Bjerg L, Dalsgaard EM, Norman K, Isaksen AA, Sandbæk A. Cohort profile: Health in Central Denmark (HICD) cohort - a register-based questionnaire survey on diabetes and related complications in the Central Denmark Region. BMJ Open. 2022;12(7):e060410. doi:10.1136/bmjopen-2021-060410

14. Schaarup JFR, Hulman A. Patients’ perception of digital technologies and artificial intelligence: an online survey. figshare. doi:10.6084/m9.figshare.19248214.v1

15. Hulman A. Peoples’ perception of artificial intelligence and chatbots: an online survey from Health in Central Denmark. figshare. doi:10.6084/m9.figshare.27894180.v2

16. Schmidt M, Schmidt SAJ, Adelborg K, et al. The Danish health care system and epidemiological research: from health care contacts to database records. Clin Epidemiol. 2019;11:563–591. doi:10.2147/CLEP.S179083

17. Pedersen CB. The Danish Civil Registration System. Scand J Public Health. 2011;39(7 Suppl):22–25. doi:10.1177/1403494810387965

18. Jensen VM, Rasmussen AW. Danish Education Registers. Scand J Public Health. 2011;39(7 Suppl):91–94. doi:10.1177/1403494810394715

19. Isaksen AA, Sandbæk A, Bjerg L. Validation of Register-Based Diabetes Classifiers in Danish Data. Clin Epidemiol. 2023;15:569–581. doi:10.2147/CLEP.S407019

20. Isaksen AA. Pre-specified analysis plan. figshare. doi:10.6084/m9.figshare.26048662.v1

21. R Core Team. R: A language and environment for statistical computing. Accessed November 29, 2024. https://www.r-project.org/

22. Barrett T, Dowle M, Srinivasan A, et al. data.table: Extension of ‘data.frame’. Accessed January 12, 2025. https://rdatatable.gitlab.io/data.table/

23. Greifer N. WeightIt: Weighting for Covariate Balance in Observational Studies. Accessed January 12, 2025. https://ngreifer.github.io/WeightIt/

24. Venables WN, Ripley BD. Modern Applied Statistics with S (nnet package). Accessed January 12, 2025. https://cran.r-project.org/web/packages/nnet

25. Brunson JC. ggalluvial: Layered Grammar for Alluvial Plots. J Open Source Softw. 2020;5(49):2017. doi:10.21105/joss.02017

26. Thomas Lin Pedersen. patchwork: The Composer of Plots. Accessed January 12, 2025. https://patchwork.data-imaginist.com/

27. Public attitudes to data and AI: Tracker survey (Wave 3). Accessed December 13, 2024. https://www.gov.uk/government/publications/public-attitudes-to-data-and-ai-tracker-survey-wave-3/public-attitudes-to-data-and-ai-tracker-survey-wave-3

28. Cresswell KM, Bates DW, Sheikh A. Ten key considerations for the successful implementation and adoption of large-scale health information technology. J Am Med Inform Assoc JAMIA. 2013;20(e1):e9–e13. doi:10.1136/amiajnl-2013-001684

29. Hoppe BF, Rueckel J, Dikhtyar Y, et al. Implementing Artificial Intelligence for Emergency Radiology Impacts Physicians’ Knowledge and Perception: A Prospective Pre-and Post-Analysis. Invest Radiol. 2024;59(5):404–412. doi:10.1097/RLI.0000000000001034

30. Kim SH, Tae JH, Chang IH, et al. Changes in patient perceptions regarding ChatGPT-written explanations on lifestyle modifications for preventing urolithiasis recurrence. Digit Health. Published online September 28, 2023. doi:10.1177/20552076231203940

31. Li LY, Isaksen AA, Lebiecka-Johansen B, et al. Prediction of cardiovascular markers and diseases using retinal fundus images and deep learning: a systematic scoping review. Eur Heart J - Digit Health. 2024;5(6):660–669. doi:10.1093/ehjdh/ztae068

32. Lim JI, Regillo CD, Sadda SR, et al. Artificial Intelligence Detection of Diabetic Retinopathy. Ophthalmol Sci. 2022;3(1):100228. doi:10.1016/j.xops.2022.100228

33. Pauling C, Kanber B, Arthurs OJ, Shelmerdine SC. Commercially available artificial intelligence tools for fracture detection: the evidence. BJR|Open. 2023;6(1):tzad005. doi:10.1093/bjro/tzad005

34. Scholz ML, Collatz-Christensen H, Blomberg SNF, Boebel S, Verhoeven J, Krafft T. Artificial intelligence in Emergency Medical Services dispatching: assessing the potential impact of an automatic speech recognition software on stroke detection taking the Capital Region of Denmark as case in point. Scand J Trauma Resusc Emerg Med. 2022;30:36. doi:10.1186/s13049-022-01020-6

35. Tierney AA, Gayre G, Hoberman B, et al. Ambient Artificial Intelligence Scribes to Alleviate the Burden of Clinical Documentation. NEJM Catal. 2024;5(3):CAT.23.0404. doi:10.1056/CAT.23.0404

36. Almanaa M. Trends and Public Perception of Artificial Intelligence in Medical Imaging: A Social Media Analysis. Cureus. 2024;16(9):e70008. doi:10.7759/cureus.70008

37. Kung TH, Cheatham M, Medenilla A, et al. Performance of ChatGPT on USMLE: Potential for AI-assisted medical education using large language models. PLOS Digit Health. 2023;2(2):e0000198. doi:10.1371/journal.pdig.0000198

38. Brin D, Sorin V, Vaid A, et al. Comparing ChatGPT and GPT-4 performance in USMLE soft skill assessments. Sci Rep. 2023;13(1):16492. doi:10.1038/s41598-023-43436-9

39. Katz U, Cohen E, Shachar E, et al. GPT versus Resident Physicians — A Benchmark Based on Official Board Scores. NEJM AI. 2024;1(5):AIdbp2300192. doi:10.1056/AIdbp2300192

40. Tanaka Y, Nakata T, Aiga K, et al. Performance of Generative Pretrained Transformer on the National Medical Licensing Examination in Japan. PLOS Digit Health. 2024;3(1):e0000433. doi:10.1371/journal.pdig.0000433

41. Ayers JW, Poliak A, Dredze M, et al. Comparing Physician and Artificial Intelligence Chatbot Responses to Patient Questions Posted to a Public Social Media Forum. JAMA Intern Med. 2023;183(6):589–596. doi:10.1001/jamainternmed.2023.1838

42. Arvidsson R, Gunnarsson R, Entezarjou A, Sundemo D, Wikberg C. ChatGPT (GPT-4) versus doctors on complex cases of the Swedish family medicine specialist examination: an observational comparative study. BMJ Open. 2024;14(12):e086148. doi:10.1136/bmjopen-2024-086148

43. Wang J, Redelmeier DA. Cognitive Biases and Artificial Intelligence. NEJM AI. 2024;1(12):AIcs2400639. doi:10.1056/AIcs2400639

44. Choi BCK, Pak AWP. A Catalog of Biases in Questionnaires. Prev Chronic Dis. 2004;2(1):A13.

