## Supplementary material for "Changes in public perception of AI in healthcare after exposure to ChatGPT"

S1: Table: Study population characteristics by ChatGPT exposure status, weighted to background population by sex, age, education level and diabetes status.

|  |  | Overall | ChatGPT exposed | No ChatGPT exposure |
| --- | --- | --- | --- | --- |
| Total N (%) |  | 5899 | 1468 (24.9) | 4431 (75.1) |
| Sex | Men | 2948 (50.0) | 791 (53.9) | 2157 (48.7) |
| Age (years) | Mean (SD) | 60.7 (10.3) | 55.9 (10.6) | 62.3 (9.7) |
| Diabetes status | No diabetes | 5095 (86.4) | 1294 (88.1) | 3801 (85.8) |
|  | Type 1 diabetes | 88 (1.5) | 30 (2.1) | 57 (1.3) |
|  | Type 2 diabetes | 716 (12.1) | 144 (9.8) | 572 (12.9) |
| Education level (years) | < 10 | 474 (8.0) | 37 (2.5) | 437 (9.9) |
|  | 10 - 15 | 3441 (58.3) | 689 (46.9) | 2752 (62.1) |
|  | > 15 | 1984 (33.6) | 742 (50.6) | 1242 (28.0) |
| Perception of AI at baseline | Don't know | 2230 (37.8) | 297 (20.2) | 1933 (43.6) |
|  | Benefits | 2325 (39.4) | 882 (60.1) | 1443 (32.6) |
|  | Equal | 1128 (19.1) | 245 (16.7) | 883 (19.9) |
|  | Risks | 216 (3.7) | 44 (3.0) | 172 (3.9) |
| Perception of AI at follow-up | Don't know | 2260 (38.3) | 260 (17.7) | 2000 (45.1) |
|  | Benefits | 2023 (34.3) | 795 (54.2) | 1228 (27.7) |
|  | Equal | 1204 (20.4) | 298 (20.3) | 905 (20.4) |
|  | Risks | 412 (7.0) | 114 (7.8) | 298 (6.7) |

S2: Table: Study population characteristics by baseline perception of AI

|  | Baseline perception | Don't know | Benefits | Equal | Risks |
| --- | --- | --- | --- | --- | --- |
| Total N (%) |  | 2236 (37.9) | 2384 (40.4) | 1083 (18.4) | 196 (3.3) |
| Sex | Men | 1069 (47.8) | 1782 (74.7) | 676 (62.4) | 122 (62.2) |
| Age (years) | Mean (SD) | 66.4 (8.3) | 64.6 (9.1) | 65.2 (9.3) | 64.9 (8.7) |
| Diabetes status | No diabetes | 1073 (48.0) | 1260 (52.9) | 558 (51.5) | 103 (52.6) |
|  | Type 1 diabetes | 131 (5.9) | 163 (6.8) | 59 (5.4) | 12 (6.1) |
|  | Type 2 diabetes | 1032 (46.2) | 961 (40.3) | 466 (43.0) | 81 (41.3) |
| Education level (years) | < 10 | 172 (7.7) | 48 (2.0) | 57 (5.3) | 12 (6.1) |
|  | 10 - 15 | 1433 (64.1) | 1162 (48.7) | 622 (57.4) | 117 (59.7) |
|  | > 15 | 631 (28.2) | 1174 (49.2) | 404 (37.3) | 67 (34.2) |
| Perception of AI at follow-up | Don't know | 1396 (62.4) | 492 (20.6) | 364 (33.6) | 73 (37.2) |
|  | Benefits | 337 (15.1) | 1385 (58.1) | 285 (26.3) | 37 (18.9) |
|  | Equal | 339 (15.2) | 400 (16.8) | 352 (32.5) | 41 (20.9) |
|  | Risks | 164 (7.3) | 107 (4.5) | 82 (7.6) | 45 (23.0) |
| ChatGPT use | Exposed | 226 (10.1) | 735 (30.8) | 202 (18.7) | 32 (16.3) |

S3: Figure: Unweighted analyses

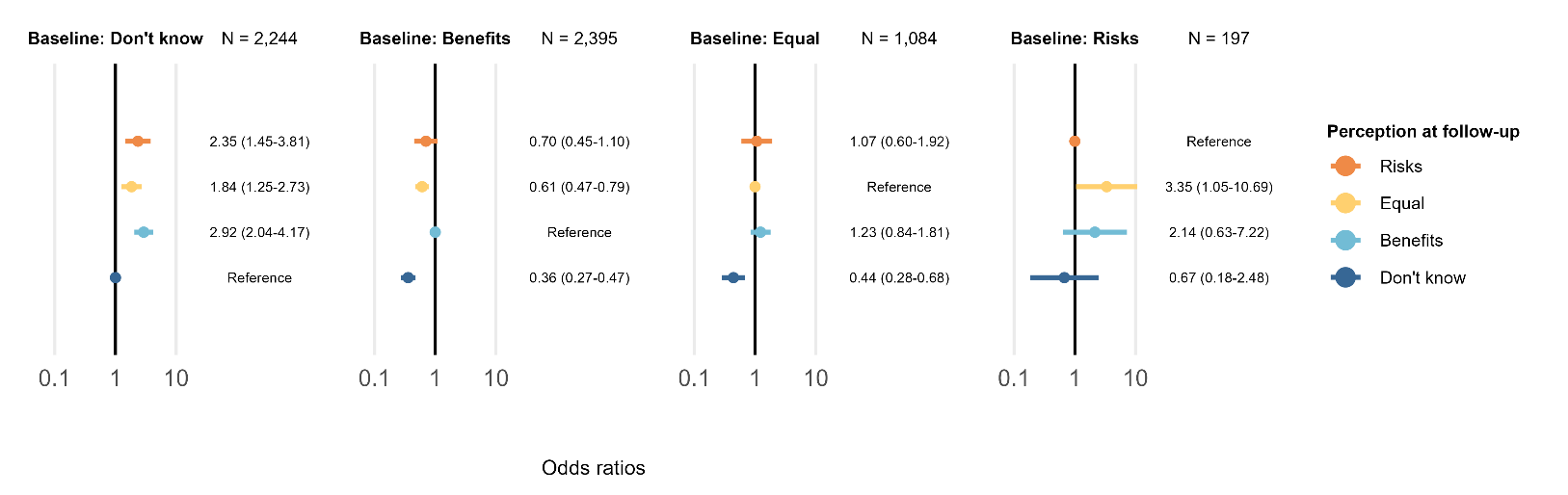

S4: Figure: Weighted to original Health in Central Denmark invitee population:

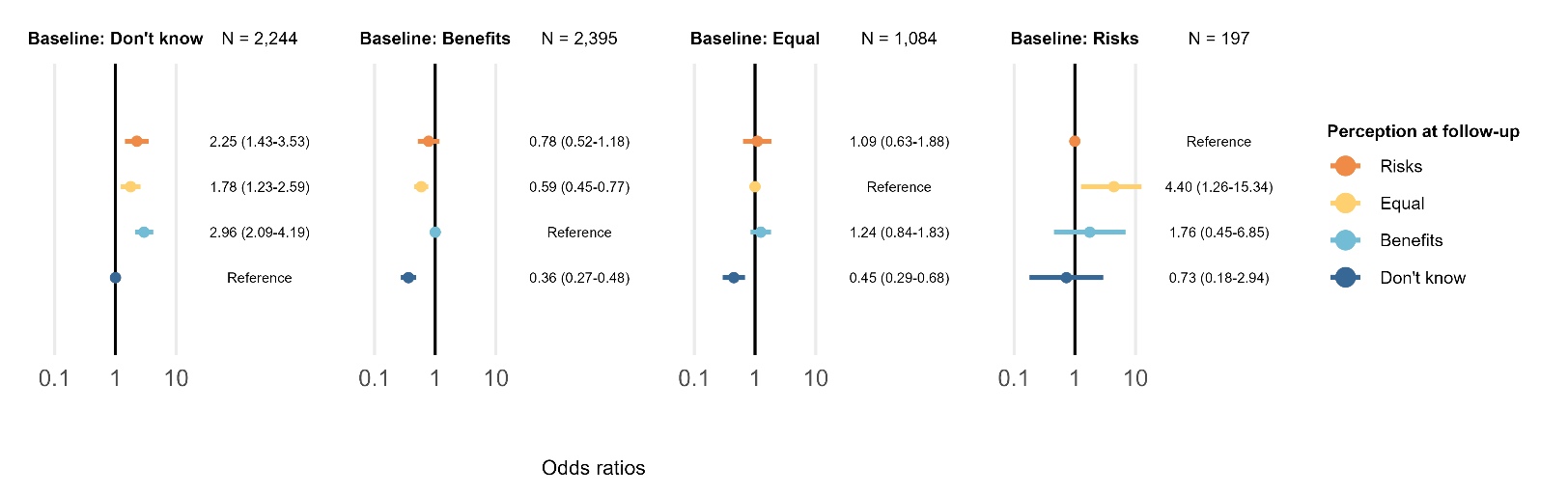

S5: Table: Full list of model coefficients.

| Baseline perception | Follow-up perception | Variable: value | Estimate (95% CI) |
| --- | --- | --- | --- |
| Don't know | Benefits | (Intercept) | 0.24 (0.08-0.76) |
| Don't know | Benefits | ChatGPT use: exposed | 3.21 (2.34-4.40) |
| Don't know | Benefits | Sex: Women | 0.43 (0.33-0.57) |
| Don't know | Benefits | Age: Per 10 years | 0.86 (0.75-0.98) |
| Don't know | Benefits | Diabetes: Type 1 | 1.30 (0.44-3.82) |
| Don't know | Benefits | Diabetes: Type 2 | 0.82 (0.55-1.23) |
| Don't know | Benefits | Education level: 10-15 | 3.01 (1.68-5.39) |
| Don't know | Benefits | Education level: >15 | 8.03 (4.36-14.80) |
| Don't know | Equal | (Intercept) | 0.21 (0.07-0.62) |
| Don't know | Equal | ChatGPT use: exposed | 1.46 (1.01-2.11) |
| Don't know | Equal | Sex: Women | 0.80 (0.62-1.04) |
| Don't know | Equal | Age: Per 10 years | 0.98 (0.85-1.12) |
| Don't know | Equal | Diabetes: Type 1 | 1.73 (0.63-4.77) |
| Don't know | Equal | Diabetes: Type 2 | 1.19 (0.84-1.67) |
| Don't know | Equal | Education level: 10-15 | 1.34 (0.90-1.99) |
| Don't know | Equal | Education level: >15 | 2.15 (1.37-3.37) |
| Don't know | Risks | (Intercept) | 0.28 (0.07-1.24) |
| Don't know | Risks | ChatGPT use: exposed | 1.40 (0.84-2.32) |
| Don't know | Risks | Sex: Women | 0.84 (0.58-1.22) |
| Don't know | Risks | Age: Per 10 years | 0.79 (0.66-0.95) |
| Don't know | Risks | Diabetes: Type 1 | 1.01 (0.20-5.04) |
| Don't know | Risks | Diabetes: Type 2 | 1.65 (1.07-2.54) |
| Don't know | Risks | Education level: 10-15 | 1.67 (0.90-3.10) |
| Don't know | Risks | Education level: >15 | 1.68 (0.82-3.44) |
| Benefits | Don't know | (Intercept) | 0.13 (0.04-0.39) |
| Benefits | Don't know | ChatGPT use: exposed | 0.32 (0.24-0.42) |
| Benefits | Don't know | Sex: Women | 2.24 (1.77-2.84) |
| Benefits | Don't know | Age: Per 10 years | 1.14 (1.00-1.29) |
| Benefits | Don't know | Diabetes: Type 1 | 1.68 (0.69-4.12) |
| Benefits | Don't know | Diabetes: Type 2 | 1.32 (0.93-1.89) |
| Benefits | Don't know | Education level: 10-15 | 1.38 (0.79-2.41) |
| Benefits | Don't know | Education level: >15 | 0.80 (0.45-1.43) |
| Benefits | Equal | (Intercept) | 0.33 (0.10-1.06) |
| Benefits | Equal | ChatGPT use: exposed | 0.53 (0.41-0.68) |
| Benefits | Equal | Sex: Women | 1.88 (1.49-2.37) |
| Benefits | Equal | Age: Per 10 years | 0.86 (0.77-0.97) |
| Benefits | Equal | Diabetes: Type 1 | 1.22 (0.54-2.74) |
| Benefits | Equal | Diabetes: Type 2 | 1.10 (0.76-1.58) |
| Benefits | Equal | Education level: 10-15 | 2.78 (1.24-6.26) |
| Benefits | Equal | Education level: >15 | 1.77 (0.78-4.04) |
| Benefits | Risks | (Intercept) | 0.62 (0.10-3.84) |
| Benefits | Risks | ChatGPT use: exposed | 0.96 (0.63-1.44) |
| Benefits | Risks | Sex: Women | 1.15 (0.75-1.77) |
| Benefits | Risks | Age: Per 10 years | 0.71 (0.58-0.86) |
| Benefits | Risks | Diabetes: Type 1 | 2.02 (0.72-5.67) |
| Benefits | Risks | Diabetes: Type 2 | 1.55 (0.90-2.70) |
| Benefits | Risks | Education level: 10-15 | 1.28 (0.40-4.06) |
| Benefits | Risks | Education level: >15 | 0.53 (0.16-1.77) |
| Equal | Benefits | (Intercept) | 0.10 (0.03-0.38) |
| Equal | Benefits | ChatGPT use: exposed | 1.02 (0.70-1.48) |
| Equal | Benefits | Sex: Women | 0.82 (0.59-1.14) |
| Equal | Benefits | Age: Per 10 years | 1.24 (1.06-1.46) |
| Equal | Benefits | Diabetes: Type 1 | 3.20 (0.61-16.72) |
| Equal | Benefits | Diabetes: Type 2 | 1.42 (0.85-2.37) |
| Equal | Benefits | Education level: 10-15 | 1.65 (0.83-3.31) |
| Equal | Benefits | Education level: >15 | 2.53 (1.21-5.28) |
| Equal | Don't know | (Intercept) | 0.59 (0.19-1.87) |
| Equal | Don't know | ChatGPT use: exposed | 0.47 (0.32-0.69) |
| Equal | Don't know | Sex: Women | 1.18 (0.87-1.59) |
| Equal | Don't know | Age: Per 10 years | 1.09 (0.95-1.26) |
| Equal | Don't know | Diabetes: Type 1 | 3.13 (0.70-14.01) |
| Equal | Don't know | Diabetes: Type 2 | 1.61 (1.03-2.54) |
| Equal | Don't know | Education level: 10-15 | 0.96 (0.55-1.67) |
| Equal | Don't know | Education level: >15 | 0.79 (0.43-1.47) |
| Equal | Risks | (Intercept) | 2.24 (0.47-10.60) |
| Equal | Risks | ChatGPT use: exposed | 0.94 (0.57-1.54) |
| Equal | Risks | Sex: Women | 0.88 (0.56-1.37) |
| Equal | Risks | Age: Per 10 years | 0.73 (0.61-0.88) |
| Equal | Risks | Diabetes: Type 1 | 1.95 (0.26-14.68) |
| Equal | Risks | Diabetes: Type 2 | 1.21 (0.61-2.43) |
| Equal | Risks | Education level: 10-15 | 0.64 (0.26-1.60) |
| Equal | Risks | Education level: >15 | 1.09 (0.41-2.84) |
| Risks | Equal | (Intercept) | 0.50 (0.00-74.74) |
| Risks | Equal | ChatGPT use: exposed | 1.40 (0.51-3.85) |
| Risks | Equal | Sex: Women | 1.12 (0.47-2.70) |
| Risks | Equal | Age: Per 10 years | 0.79 (0.45-1.39) |
| Risks | Equal | Diabetes: Type 1 | 0.12 (0.00-24.43) |
| Risks | Equal | Diabetes: Type 2 | 0.88 (0.23-3.33) |
| Risks | Equal | Education level: 10-15 | 16.33 (0.75-355.85) |
| Risks | Equal | Education level: >15 | 6.35 (0.27-148.94) |
| Risks | Benefits | (Intercept) | 721.01 (9.46-54966.17) |
| Risks | Benefits | ChatGPT use: exposed | 0.71 (0.24-2.10) |
| Risks | Benefits | Sex: Women | 0.36 (0.14-0.91) |
| Risks | Benefits | Age: Per 10 years | 0.38 (0.21-0.68) |
| Risks | Benefits | Diabetes: Type 1 | 0.02 (0.00-15.77) |
| Risks | Benefits | Diabetes: Type 2 | 0.21 (0.04-1.18) |
| Risks | Benefits | Education level: 10-15 | 1.68 (0.33-8.63) |
| Risks | Benefits | Education level: >15 | 0.96 (0.16-5.56) |
| Risks | Don't know | (Intercept) | 7.59 (0.13-451.71) |
| Risks | Don't know | ChatGPT use: exposed | 0.27 (0.08-0.98) |
| Risks | Don't know | Sex: Women | 1.55 (0.65-3.67) |
| Risks | Don't know | Age: Per 10 years | 0.77 (0.44-1.33) |
| Risks | Don't know | Diabetes: Type 1 | 0.73 (0.03-20.67) |
| Risks | Don't know | Diabetes: Type 2 | 0.94 (0.28-3.17) |
| Risks | Don't know | Education level: 10-15 | 2.11 (0.53-8.48) |
| Risks | Don't know | Education level: >15 | 0.30 (0.06-1.53) |

S6: Table: Unstratified multinomial regression model additionally adjusted for baseline perception. Estimates for the association between ChatGPT exposure and perception at follow-up, with “Benefits” at follow-up as the reference outcome level.

| Perception at follow-up | Estimate (95% CI) |
| --- | --- |
| Don't know | 0.35 (0.29-0.42) |
| Equal | 0.64 (0.54-0.76) |
| Risks | 0.74 (0.58-0.96) |

S7 File: PubMed search string for existing literature on the subject

As of 1 Dec 2024, this search query retrieves 12 results, none of which are relevant to the research question:

("artificial intelligence"[Title/Abstract] OR "machine learning"[Title/Abstract]) AND perception[Title/Abstract] AND (population[Title/Abstract] OR public[Title/Abstract] OR people[Title/Abstract]) AND ("follow-up"[Title/Abstract] OR "longitudinal"[Title/Abstract])
